# Effects of transcranial ultrasound stimulation on stroke recovery: a systematic review and meta-analysis of preclinical and clinical evidence

**DOI:** 10.1101/2025.04.21.25326124

**Authors:** Yueyeu Zhang, Anirban Dutta, Abhijit Das, Atif Shahzad

## Abstract

**Objective:** This systematic review and meta-analysis aim to summarize and analyze the available evidence of non-invasive transcranial ultrasound stimulation on stroke recovery in stroke patients and stroke animal models.

**Data Sources:** PubMed, Scopus, Web of Science and Cochrane Library databases were searched for relevant articles from database inception to October 31, 2024.

**Study Selection:** The inclusion criteria were randomized controlled human trials and controlled animal studies, which included trials exploring the transcranial ultrasound stimulation effects toward clinical and pre-clinical, neuroimaging, neurophysiological, and biochemical outcomes. The research identified 463 potential articles with 5 human studies and 13 animal studies included.

**Result:** The data indicates that transcranial ultrasound stimulation is effective in supporting stroke recovery, especially the motor functions. However, the results are inhomogeneous across studies: (1) transcranial ultrasound stimulation induce greater effects if applied together with other therapies, and (2) apply transcranial ultrasound stimulation at different time delay or different therapy time induce different effects on functional motor ability.

**Conclusion:** Evidence suggests that transcranial ultrasound stimulation is a promising intervention in improving daily lives of stroke patients or animals. While further research is encouraged to prove the effect sizes in stroke patients.

## 1. Introduction

Stroke is a worldwide disease caused by blood flowing into brain tissue. In 2021, Stroke was the third common cause of death and fourth cause of disability-adjusted life-years, and there was research indicated that there were about 93.8 million prevalent, 11.9 million incident strokes, and 160.5 million DALYs from stroke [1]. Some sequels of stroke will greatly affect patients’ quality of life; therefore, interventions are needed to improve recovery. Evidence shows that brain try to repair itself immediately after stroke, and there is a critical period when rehabilitation tends to work better to help people recover. This window is usually within 4.5 hours after stroke[17]. While not all patients can receive prompt treatments in acute-phase, and stroke causes irreversible neurons death and brain damage to survivors [2]. In chronic phase, the unaffected hemisphere will gain more excitability after stroke, aiming to compensate for lost function [3]. Thus, in order to reopen that window for rehabilitation, it is of great importance to modify brain excitability [4]. In consequence, two potential strategies for modulating plasticity are invasive brain stimulation and non-invasive brain stimulation. While taking into consideration that invasive stimulation may cause some side effects like pain, non-invasive ones seem to be more suitable for everyday treatment. When compared to other non-invasive approaches such as transcranial magnetic stimulation and transcranial electrical stimulation, ultrasound stimulation offers better resolution, relatively low cost and effective targeting [5]. Moreover, ultrasound stimulation is proved to have ability of neuromodulation [6], which might be applied in some stroke therapies.

The mechanism of transcranial ultrasound to neuromodulation can be explained as affecting the discharge ability of cells [7] or signaling pathway [8], then more tissue recovery related proteins are produced to help brain to form blood vessels for further repairing. Moreover, interhemispheric neuronal activities are enhanced and contribute to motor functions recovery [9]. Although there might be more than two mechanisms, ultrasound neuromodulation is considered as a new but promising brain therapy [6]. There are five main parameters that may influence the effect of transcranial ultrasound stimulation, which are frequency, peak intensity, duration, pulse repetition frequency, and duty cycle [10].

Over the years, there has been lots of research proving the effectiveness of transcranial ultrasound stimulation on stroke recovery. While the fact is that, up to now, few review studies have been done on that, and more studies are needed. This systematic review and meta-analysis aim to collect and analyze data to prove that transcranial ultrasound stimulation is a powerful tool in stroke treatment in both animal and human studies. In this paper, TUS is applied as an abbreviation of transcranial ultrasound stimulation.

## 2. Method

This systematic review and meta-analysis use the protocol that is based on the Preferred Reporting Items for Systematic Reviews and Meta-Analysis (PRISMA) guidelines and was registered on PROSPERO (CRD420250617123).

### 2.1 Search protocol

Relevant studies were identified by searching of online databases, which were PubMed, Cochrane Library, Scopus and Web of Science from inception to 31^st^ October 2024. The search terms used in these databases were revised and the language was limited to English. The following keywords were searched: “ultrasound stimulation”, “stroke” and “recovery”. The complete search strategies are presented: web of science (ultrasound stimulation and stroke recovery), PubMed (“ultrasound” and “stimulation” and stroke), Scopus (“ultrasound stimulation” and stroke recovery), Cochrane Library (ultrasound stimulation and stroke). Reference lists of relevant reviews and included studies were searched to identify potential records.

### 2.2 Eligibility criteria

The inclusion criteria for this study were as follows: (1) randomized controlled trials, (2) intervention groups receiving TUS alone or in combination with other therapies, and control groups receiving sham or no TUS, (3) measured outcomes relating to stroke recovery, (4) written in English.

The exclusion criteria were as follows: (1) studies that did not meet the inclusion criteria, (2) studies published in the form of conference abstracts, reviews and study protocols.

### 2.3 Data extraction

The data related to stroke recovery were extracted from the included publications. From each study, information obtained including the name of the first author, year of publication, number of participants, characteristics of subjects or animal models, treatment parameters and outcome measures.

### 2.4 Data synthesis

Based on the methodological approach, the experiments included were experiments comparing real brain stimulation with sham stimulation. Within this framework, a subcategorization was done depending on different experimental subjects, including tests on stroke patients, mice and healthy people.

### 2.5 Methodological quality assessment

11-items PEDro scale was applied to evaluate the methodological quality of the human studies, such as random allocation, subjects and assessors blinding, dropout rate etc. [11]. In order to evaluate animal tests, we updated some elements in previously used quality scales [12,13]. The “1” score meant that this element was clearly mentioned in the study or could be logically inferred through description; if one element was clearly not included in the study, it would gain “0” score. While if one element was not mentioned and could’t be simply inferred to Yes or No, a question mark “?” would be used for this uncertainty, and this would not be included in the final scores. In addition, if the outcomes were assessed by machines and researchers only collected the data presented by machines, it would gain “1” score. The higher the total score, the higher the methodological quality (10–9 excellent, 8–6 good, 5–4 fair and <4 poor). The RoB2 tool was additionally applied for human studies [42].

### 2.6 Strategy for qualitative synthesis

Some parts of the included studies were not easy to perform meta-analyses if considering the high heterogeneity of the included studies [14]. For the outcomes could be mainly divided into motor function recovery and cognitive rehabilitation, and they couldn’t be combined. Studies with the same outcome of interest were grouped, and the comparative results between the experimental group and the control group were summarized.

### 2.7 Statistical analysis

In this review, we only analyze the outcomes linked to recovery of motor functions considering the heterogeneity mentioned above. Effect size calculators were used to estimate the effect size and the 95% confidence interval for each experiment with mean values and standard divisions [37,38]. Depending on availability, the calculations are based on either means and standard deviations of pre-intervention and post-intervention, or on means and standard deviations of the difference between the baseline and post-intervention data. The Cohen’s d method was applied for calculation and the effect size was used for interpretation (d≥0.2 “small”, d≥0.5 “medium”, d≥0.8 “large”) [39,40]. There were some articles that only included P values, in this case, we applied the Fisher’s method [41]. The METAANALYSISONLINE was applied for drawing figures [43].

## 3. Result

### 3.1 Study selection and study characteristics

The original search of the databases identified 463 records, then Covidence identified 52 duplicates, after that, we checked the titles of 411 papers and excluded 358 papers. Then we divided into the abstracts of 61papers, finding there were 7 paper didn’t have the abstracts or couldn’t find the full texts. After these 7 papers were excluded, we went through the abstracts of 54 papers and finally 18 studies were retrieved for full-text review with matched theme. In total, 5 human studies and 13 animal studies were included in this systematic review. The study selection process is shown in Figure 1.

**Figure 1.**
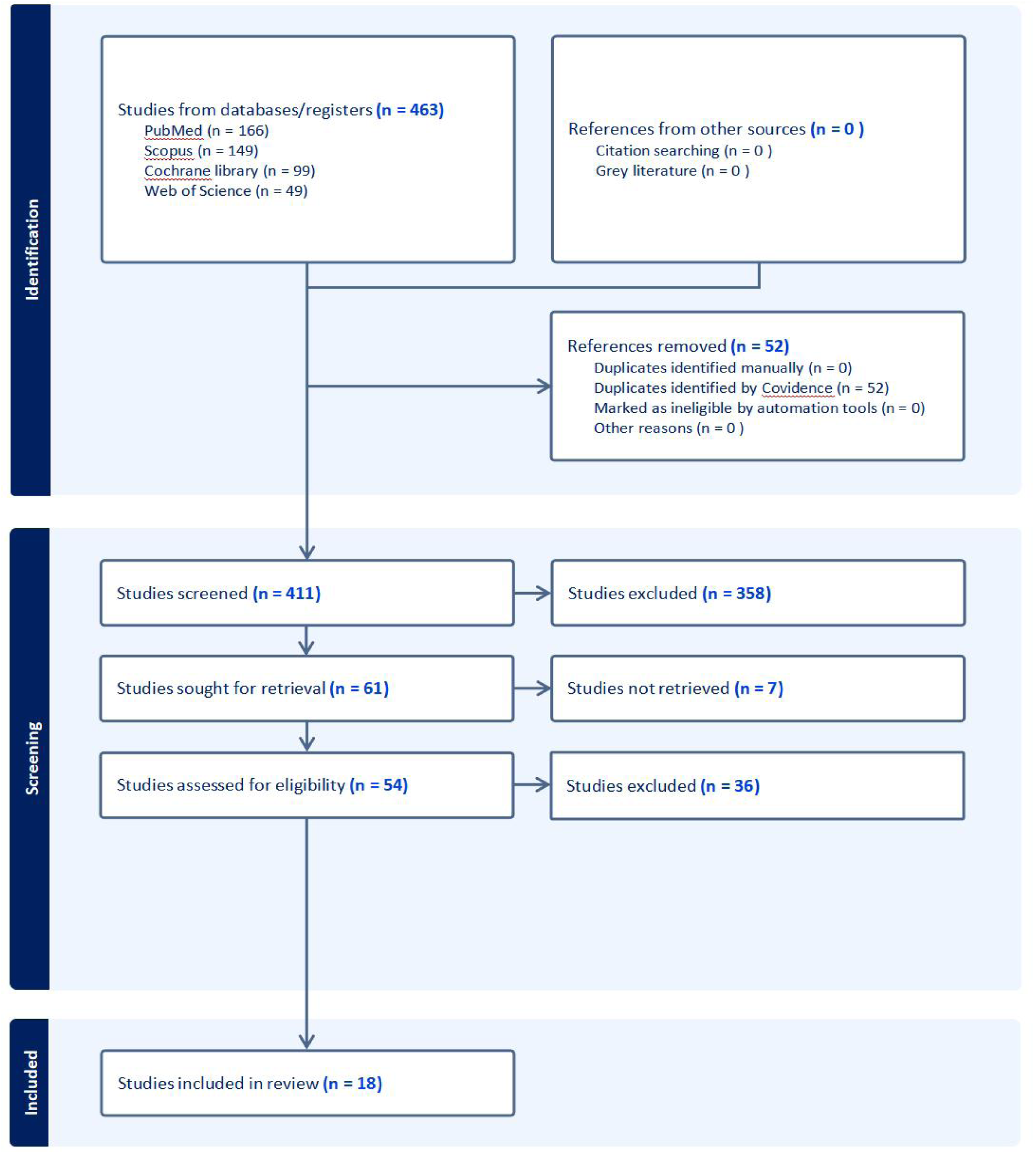
PRISMA flow diagram

Table 1 and Table 2 present detailed characteristics of individual animal and human studies, respectively. Among five human studies, three of them included healthy subjects, two studies included stroke patients. All animal studies applied mice stroke models. The group sizes of each study were not shown in these tables, for some studies didn’t divide subjects into groups, or didn’t mention the group size, in addition that some used different number of mice in different experiments.

**Table 1.**
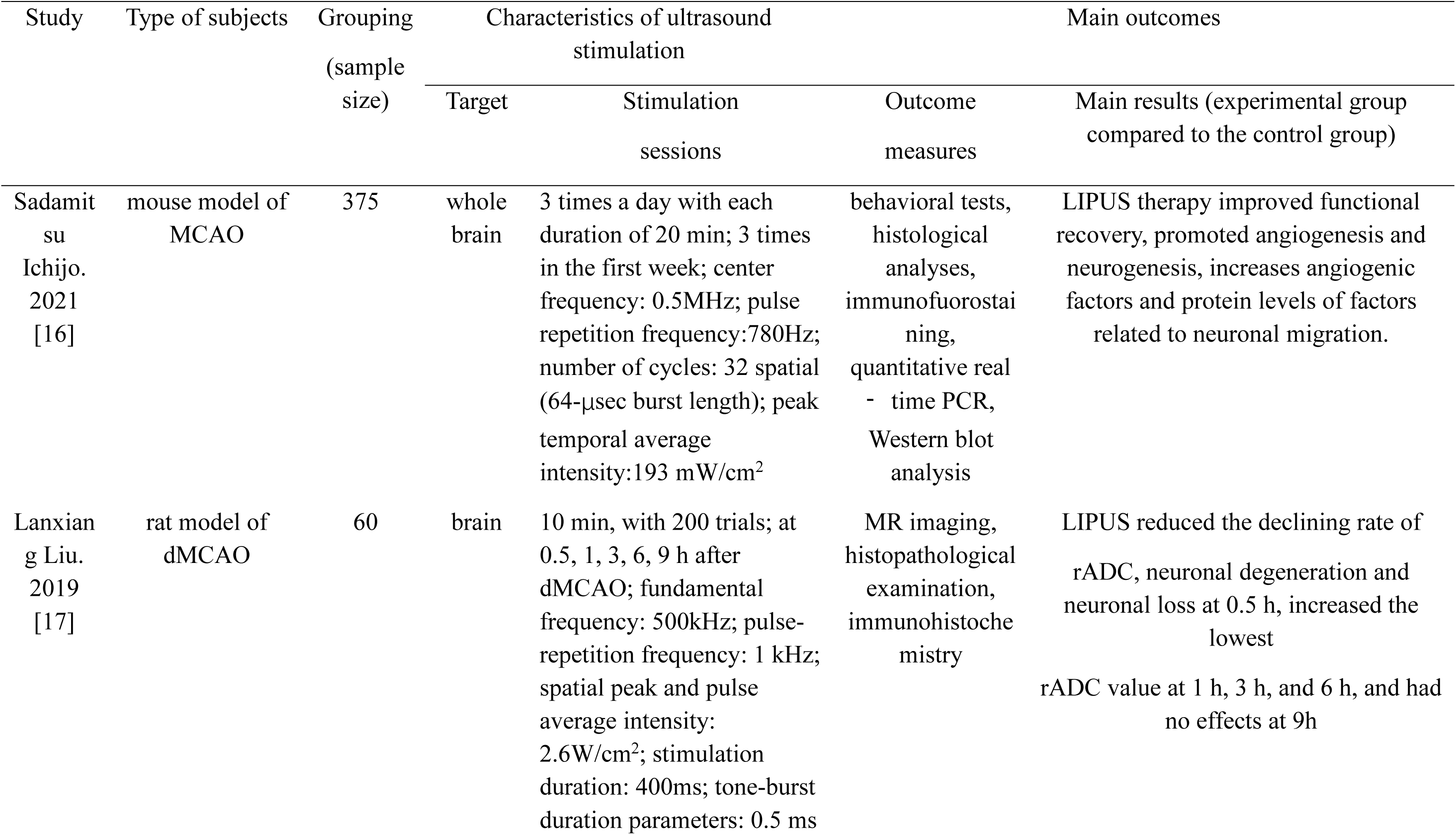
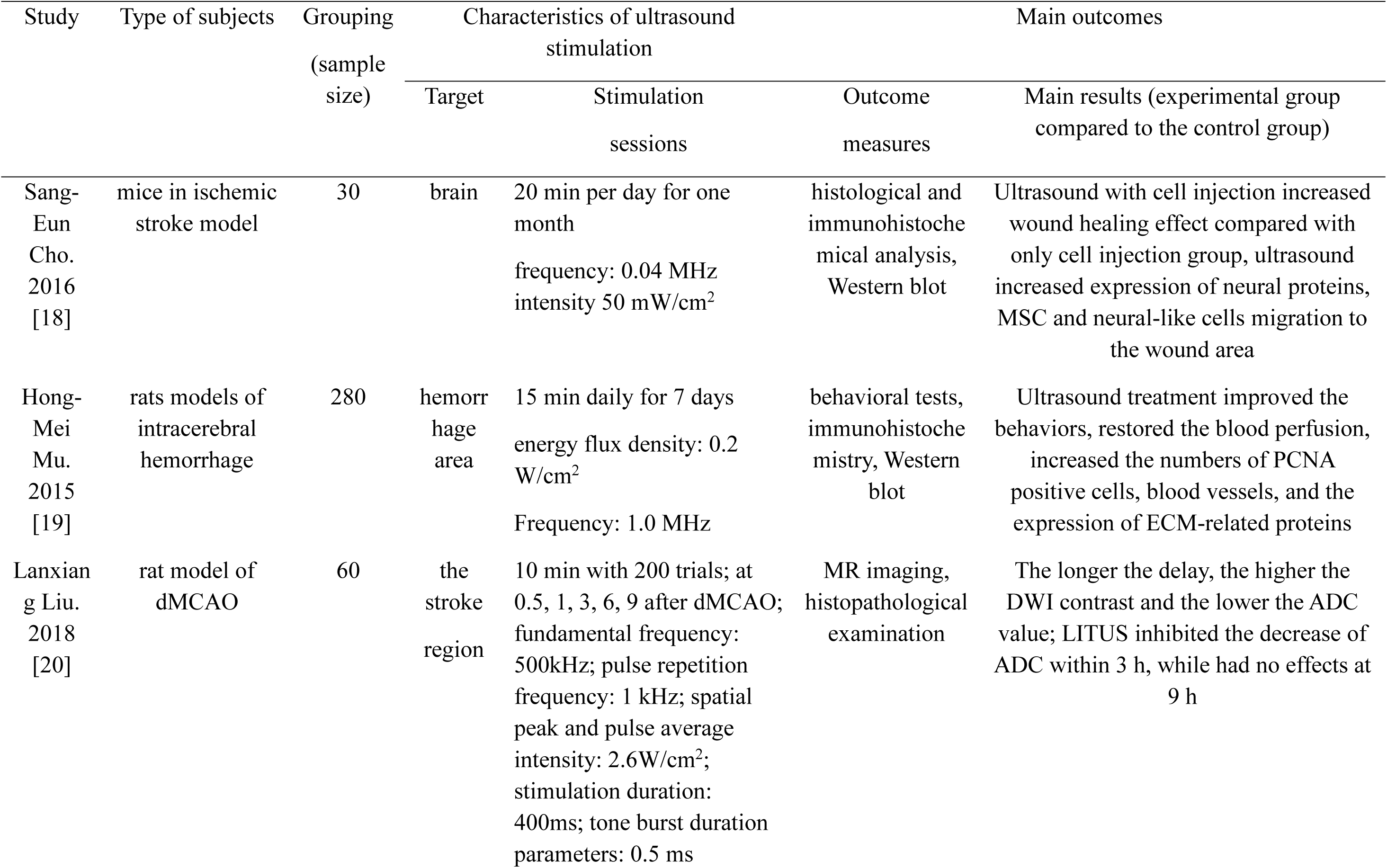
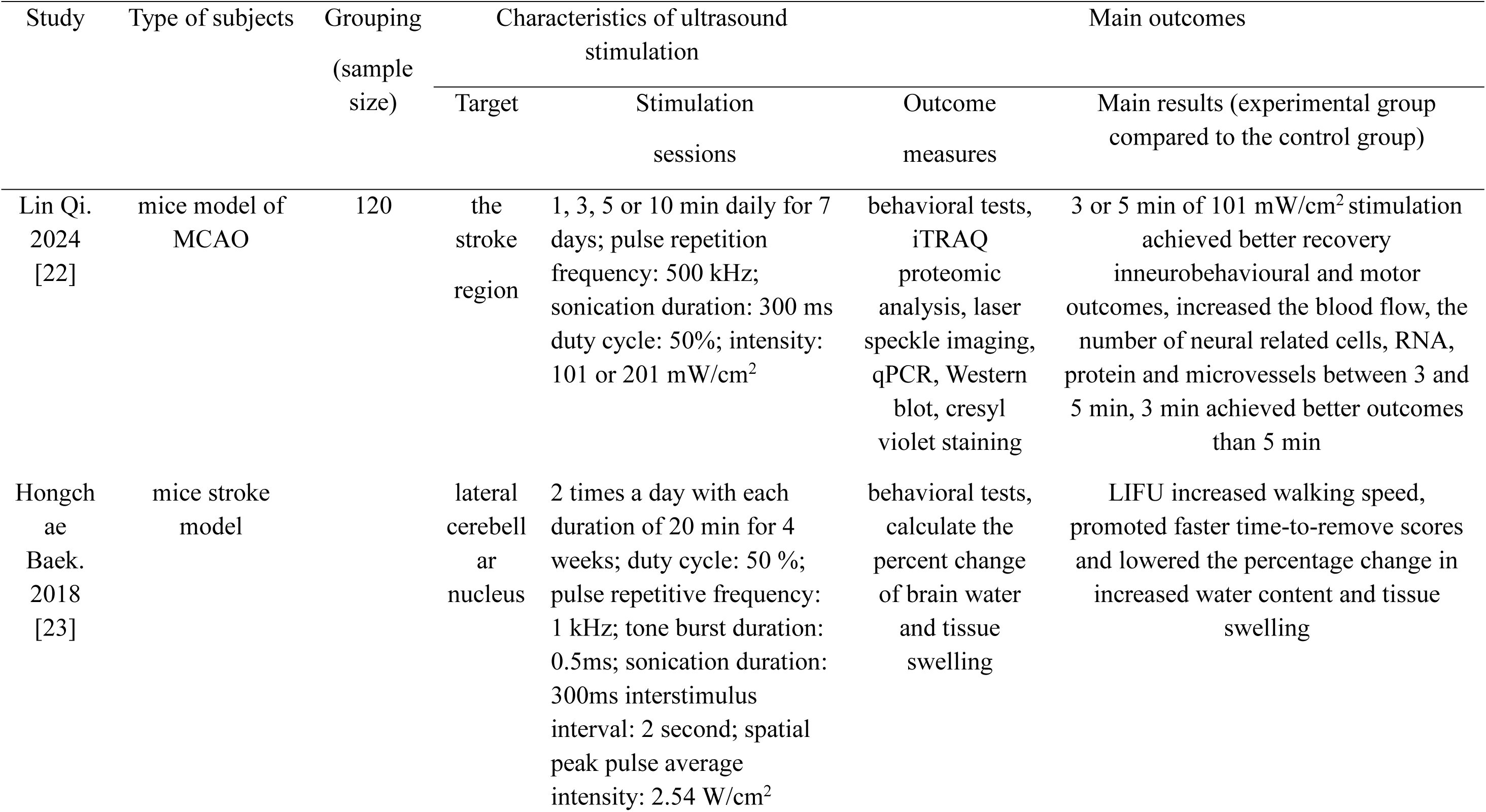
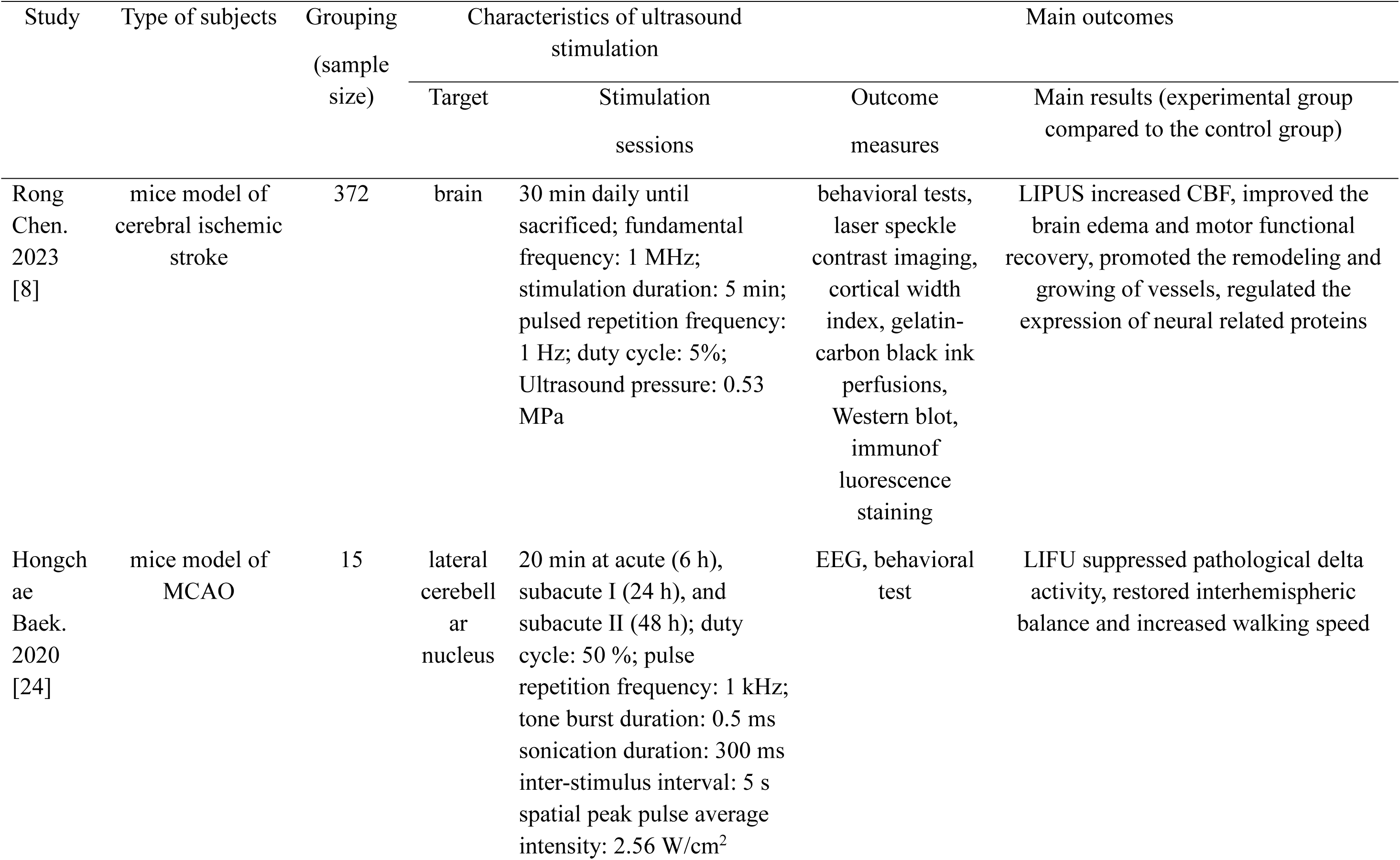
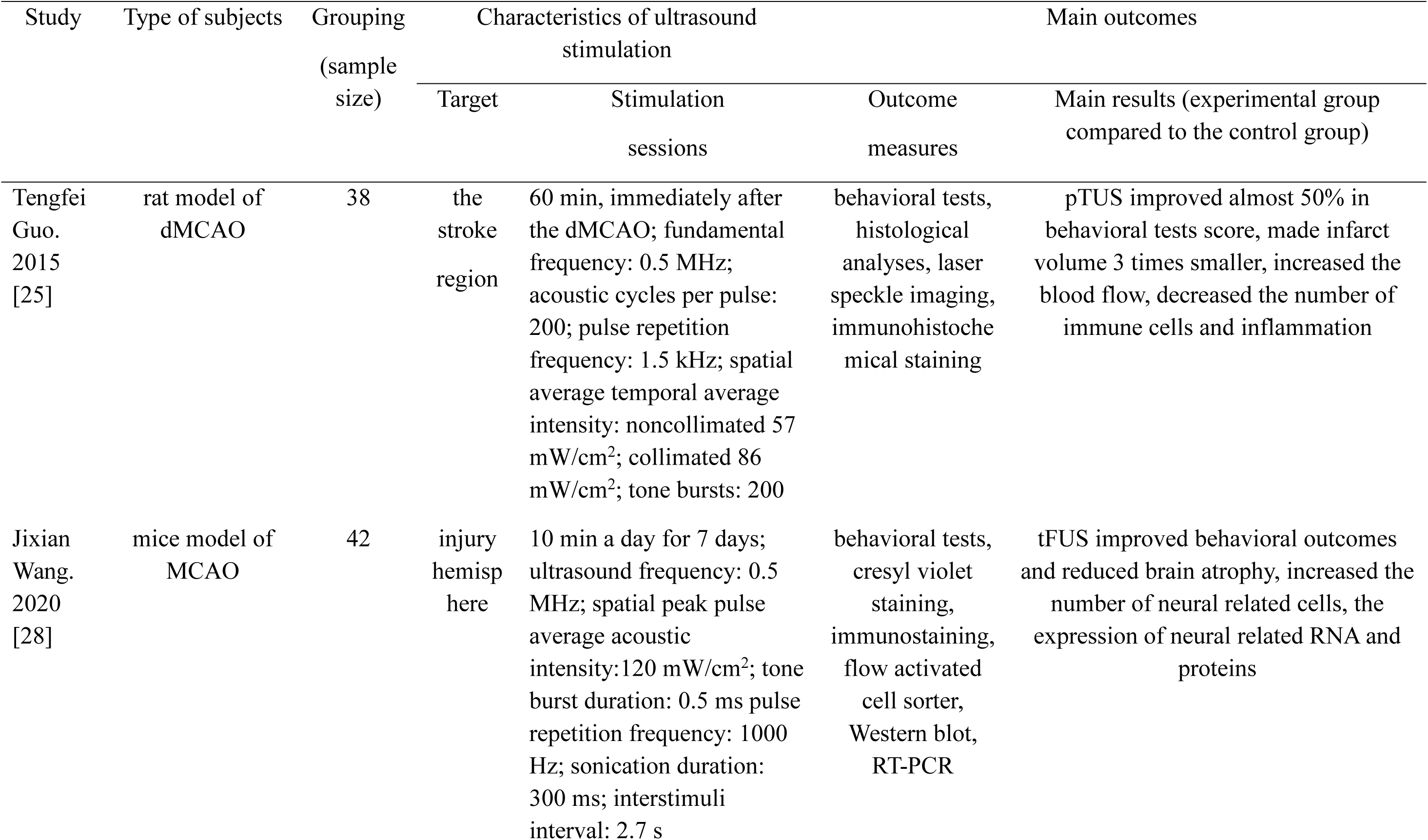
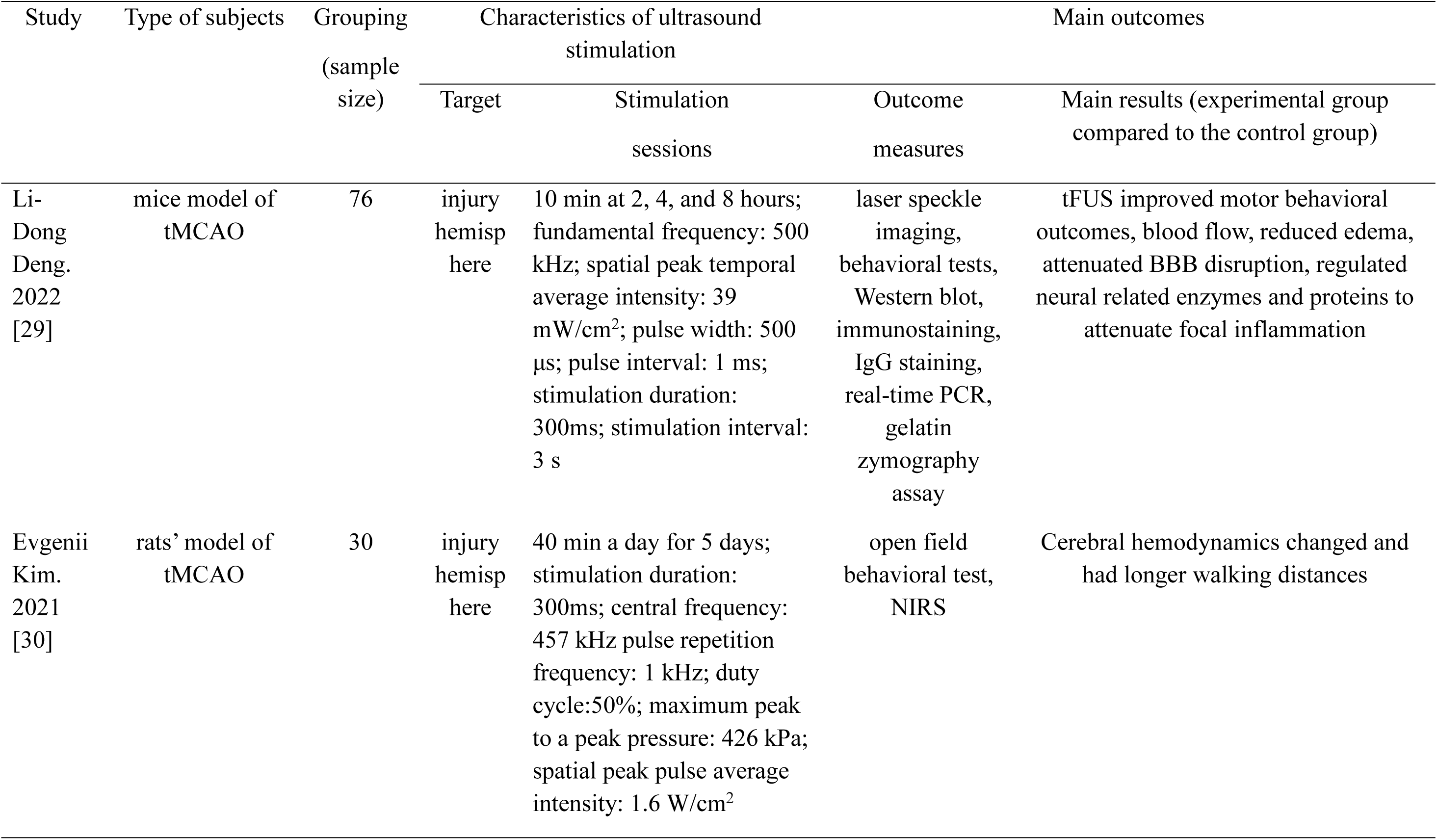
Characteristics of included animal studies.

| Study | Type of subjects | Grouping<br>(sample<br>size) | Characteristics of ultrasound<br>stimulation |  |  | Main outcomes |
| --- | --- | --- | --- | --- | --- | --- |
|  |  |  | Target | Stimulation<br>sessions | Outcome<br>measures | Main results (experimental group<br>compared to the control group) |
| Sadamitsu<br>Ichijo.<br>2021<br>[16] | mouse model of<br>MCAO | 375 | whole<br>brain | 3 times a day with each<br>duration of 20 min; 3 times<br>in the first week; center<br>frequency: 0.5MHz; pulse<br>repetition frequency:780Hz;<br>number of cycles: 32 spatial<br>(64-μsec burst length); peak<br>temporal average<br>intensity:193 mW/cm<sup>2</sup> | behavioral tests,<br>histological<br>analyses,<br>immunofluorescence,<br>quantitative real<br>- time PCR,<br>Western blot<br>analysis | LIPUS therapy improved functional<br>recovery, promoted angiogenesis and<br>neurogenesis, increases angiogenic<br>factors and protein levels of factors<br>related to neuronal migration. |
| Lanxiang<br>Liu.<br>2019<br>[17] | rat model of<br>dMCAO | 60 | brain | 10 min, with 200 trials; at<br>0.5, 1, 3, 6, 9 h after<br>dMCAO; fundamental<br>frequency: 500kHz; pulse-<br>repetition frequency: 1 kHz;<br>spatial peak and pulse<br>average intensity:<br>2.6W/cm<sup>2</sup>; stimulation<br>duration: 400ms; tone-burst<br>duration parameters: 0.5 ms | MR imaging,<br>histopathological<br>examination,<br>immunohistochemistry | LIPUS reduced the declining rate of<br>rADC, neuronal degeneration and<br>neuronal loss at 0.5 h, increased the<br>lowest<br>rADC value at 1 h, 3 h, and 6 h, and had<br>no effects at 9h |
|  |  |  | Target | Stimulation<br>sessions | Outcome<br>measures | Main results (experimental group<br>compared to the control group) |
| Sang-Eun<br>Cho.<br>2016<br>[18] | mice in ischemic<br>stroke model | 30 | brain | 20 min per day for one<br>month<br>frequency: 0.04 MHz<br>intensity 50 mW/cm<sup>2</sup> | histological and<br>immunohistoche<br>mical analysis,<br>Western blot | Ultrasound with cell injection increased<br>wound healing effect compared with<br>only cell injection group, ultrasound<br>increased expression of neural proteins,<br>MSC and neural-like cells migration to<br>the wound area |
| Hong-Mei<br>Mu.<br>2015<br>[19] | rats models of<br>intracerebral<br>hemorrhage | 280 | hemorrhage<br>area | 15 min daily for 7 days<br>energy flux density: 0.2<br>W/cm<sup>2</sup><br>Frequency: 1.0 MHz | behavioral tests,<br>immunohistoche<br>mistry, Western<br>blot | Ultrasound treatment improved the<br>behaviors, restored the blood perfusion,<br>increased the numbers of PCNA<br>positive cells, blood vessels, and the<br>expression of ECM-related proteins |
| Lanxiang<br>Liu.<br>2018<br>[20] | rat model of<br>dMCAO | 60 | the<br>stroke<br>region | 10 min with 200 trials; at<br>0.5, 1, 3, 6, 9 after dMCAO;<br>fundamental frequency:<br>500kHz; pulse repetition<br>frequency: 1 kHz; spatial<br>peak and pulse average<br>intensity: 2.6W/cm<sup>2</sup>;<br>stimulation duration:<br>400ms; tone burst duration<br>parameters: 0.5 ms | MR imaging,<br>histopathological<br>examination | The longer the delay, the higher the<br>DWI contrast and the lower the ADC<br>value; LITUS inhibited the decrease of<br>ADC within 3 h, while had no effects at<br>9 h |

| Study | Type of subjects | Grouping<br>(sample size) | Characteristics of ultrasound stimulation |  |  | Main outcomes |
| --- | --- | --- | --- | --- | --- | --- |
|  |  |  | Target | Stimulation sessions | Outcome measures | Main results (experimental group compared to the control group) |
| Lin Qi. 2024 [22] | mice model of MCAO | 120 | the stroke region | 1, 3, 5 or 10 min daily for 7 days; pulse repetition frequency: 500 kHz; sonication duration: 300 ms duty cycle: 50%; intensity: 101 or 201 mW/cm<sup>2</sup> | behavioral tests, iTRAQ proteomic analysis, laser speckle imaging, qPCR, Western blot, cresyl violet staining | 3 or 5 min of 101 mW/cm<sup>2</sup> stimulation achieved better recovery in neurobehavioural and motor outcomes, increased the blood flow, the number of neural related cells, RNA, protein and microvessels between 3 and 5 min, 3 min achieved better outcomes than 5 min |
| Hongchae Baek. 2018 [23] | mice stroke model |  | lateral cerebellar nucleus | 2 times a day with each duration of 20 min for 4 weeks; duty cycle: 50 %; pulse repetitive frequency: 1 kHz; tone burst duration: 0.5ms; sonication duration: 300ms interstimulus interval: 2 second; spatial peak pulse average intensity: 2.54 W/cm<sup>2</sup> | behavioral tests, calculate the percent change of brain water and tissue swelling | LIFU increased walking speed, promoted faster time-to-remove scores and lowered the percentage change in increased water content and tissue swelling |

*(continued on next page)*

| Study | Type of subjects | Grouping<br>(sample<br>size) | Characteristics of ultrasound<br>stimulation |  |  | Main outcomes |
| --- | --- | --- | --- | --- | --- | --- |
|  |  |  | Target | Stimulation<br>sessions | Outcome<br>measures | Main results (experimental group<br>compared to the control group) |
| Rong<br>Chen.<br>2023<br>[8] | mice model of<br>cerebral ischemic<br>stroke | 372 | brain | 30 min daily until<br>sacrificed; fundamental<br>frequency: 1 MHz;<br>stimulation duration: 5 min;<br>pulsed repetition frequency:<br>1 Hz; duty cycle: 5%;<br>Ultrasound pressure: 0.53<br>MPa | behavioral tests,<br>laser speckle<br>contrast imaging,<br>cortical width<br>index, gelatin-<br>carbon black ink<br>perfusions,<br>Western blot,<br>immunof<br>luorescence<br>staining | LIPUS increased CBF, improved the<br>brain edema and motor functional<br>recovery, promoted the remodeling and<br>growing of vessels, regulated the<br>expression of neural related proteins |
| Hongch<br>ae<br>Baek.<br>2020<br>[24] | mice model of<br>MCAO | 15 | lateral<br>cerebell<br>ar<br>nucleus | 20 min at acute (6 h),<br>subacute I (24 h), and<br>subacute II (48 h); duty<br>cycle: 50 %; pulse<br>repetition frequency: 1 kHz;<br>tone burst duration: 0.5 ms<br>sonication duration: 300 ms<br>inter-stimulus interval: 5 s<br>spatial peak pulse average<br>intensity: 2.56 W/cm<sup>2</sup> | EEG, behavioral<br>test | LIFU suppressed pathological delta<br>activity, restored interhemispheric<br>balance and increased walking speed |

*(continued on next page)*

| Study | Type of subjects | Grouping<br>(sample<br>size) | Characteristics of ultrasound<br>stimulation |  |  | Main outcomes |
| --- | --- | --- | --- | --- | --- | --- |
|  |  |  | Target | Stimulation<br>sessions | Outcome<br>measures | Main results (experimental group<br>compared to the control group) |
| Tengfei<br>Guo.<br>2015<br>[25] | rat model of<br>dMCAO | 38 | the<br>stroke<br>region | 60 min, immediately after<br>the dMCAO; fundamental<br>frequency: 0.5 MHz;<br>acoustic cycles per pulse:<br>200; pulse repetition<br>frequency: 1.5 kHz; spatial<br>average temporal average<br>intensity: noncollimated 57<br>mW/cm<sup>2</sup>; collimated 86<br>mW/cm<sup>2</sup>; tone bursts: 200 | behavioral tests,<br>histological<br>analyses, laser<br>speckle imaging,<br>immunohistoche<br>mical staining | pTUS improved almost 50% in<br>behavioral tests score, made infarct<br>volume 3 times smaller, increased the<br>blood flow, decreased the number of<br>immune cells and inflammation |
| Jixian<br>Wang.<br>2020<br>[28] | mice model of<br>MCAO | 42 | injury<br>hemisp<br>here | 10 min a day for 7 days;<br>ultrasound frequency: 0.5<br>MHz; spatial peak pulse<br>average acoustic<br>intensity: 120 mW/cm<sup>2</sup>; tone<br>burst duration: 0.5 ms pulse<br>repetition frequency: 1000<br>Hz; sonication duration:<br>300 ms; interstimuli<br>interval: 2.7 s | behavioral tests,<br>cresyl violet<br>staining,<br>immunostaining,<br>flow activated<br>cell sorter,<br>Western blot,<br>RT-PCR | tFUS improved behavioral outcomes<br>and reduced brain atrophy, increased the<br>number of neural related cells, the<br>expression of neural related RNA and<br>proteins |

*(continued on next page)*

| Study | Type of subjects | Grouping<br>(sample size) | Characteristics of ultrasound stimulation |  |  | Main outcomes |
| --- | --- | --- | --- | --- | --- | --- |
|  |  |  | Target | Stimulation sessions | Outcome measures | Main results (experimental group compared to the control group) |
| Li-Dong Deng, 2022 [29] | mice model of tMCAO | 76 | injury hemisp here | 10 min at 2, 4, and 8 hours; fundamental frequency: 500 kHz; spatial peak temporal average intensity: 39 mW/cm<sup>2</sup>; pulse width: 500 µs; pulse interval: 1 ms; stimulation duration: 300ms; stimulation interval: 3 s | laser speckle imaging, behavioral tests, Western blot, immunostaining, IgG staining, real-time PCR, gelatin zymography assay | tFUS improved motor behavioral outcomes, blood flow, reduced edema, attenuated BBB disruption, regulated neural related enzymes and proteins to attenuate focal inflammation |
| Evgenii Kim, 2021 [30] | rats' model of tMCAO | 30 | injury hemisp here | 40 min a day for 5 days; stimulation duration: 300ms; central frequency: 457 kHz pulse repetition frequency: 1 kHz; duty cycle: 50%; maximum peak to a peak pressure: 426 kPa; spatial peak pulse average intensity: 1.6 W/cm<sup>2</sup> | open field behavioral test, NIRS | Cerebral hemodynamics changed and had longer walking distances |

**Table 2.**
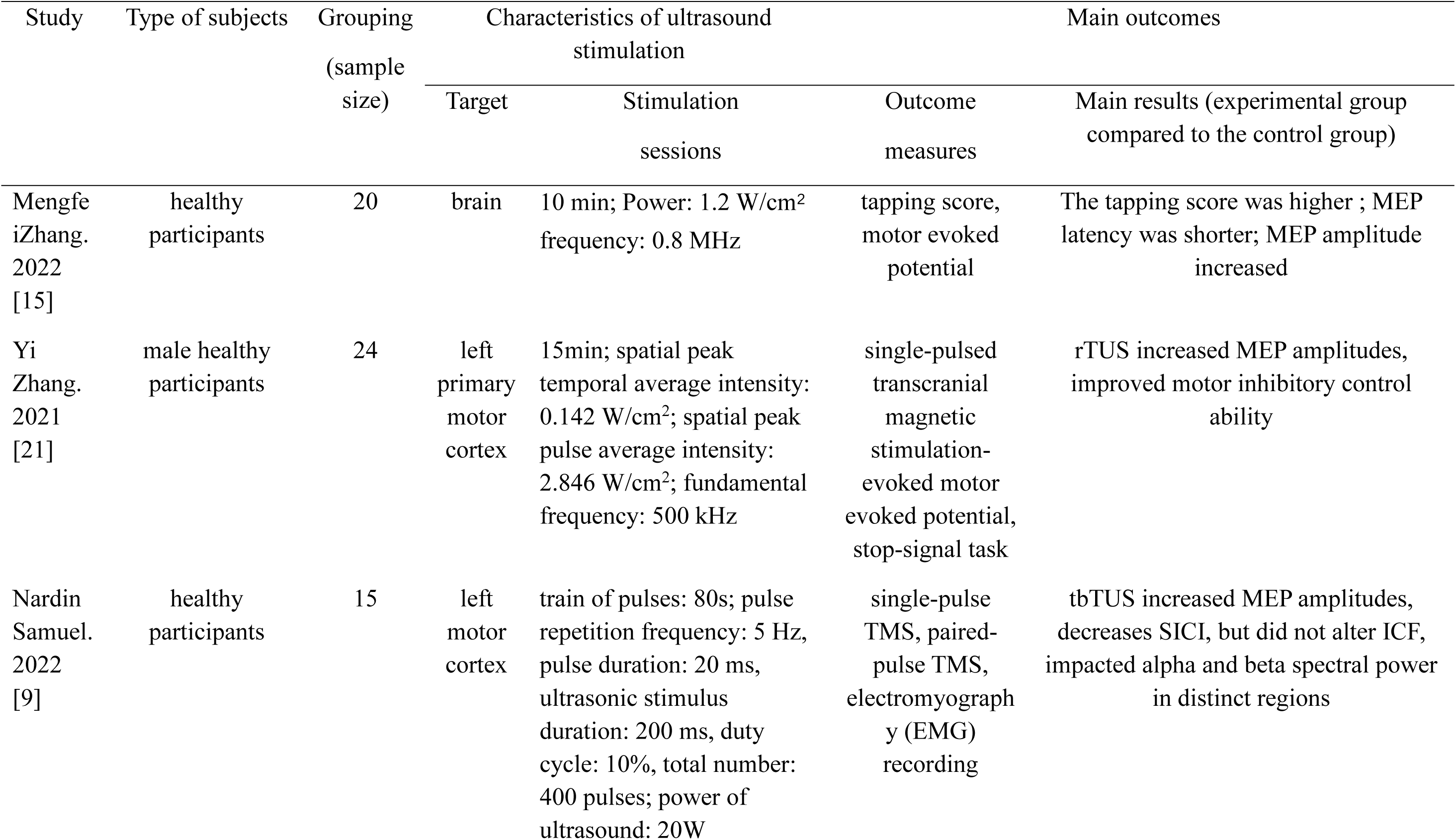
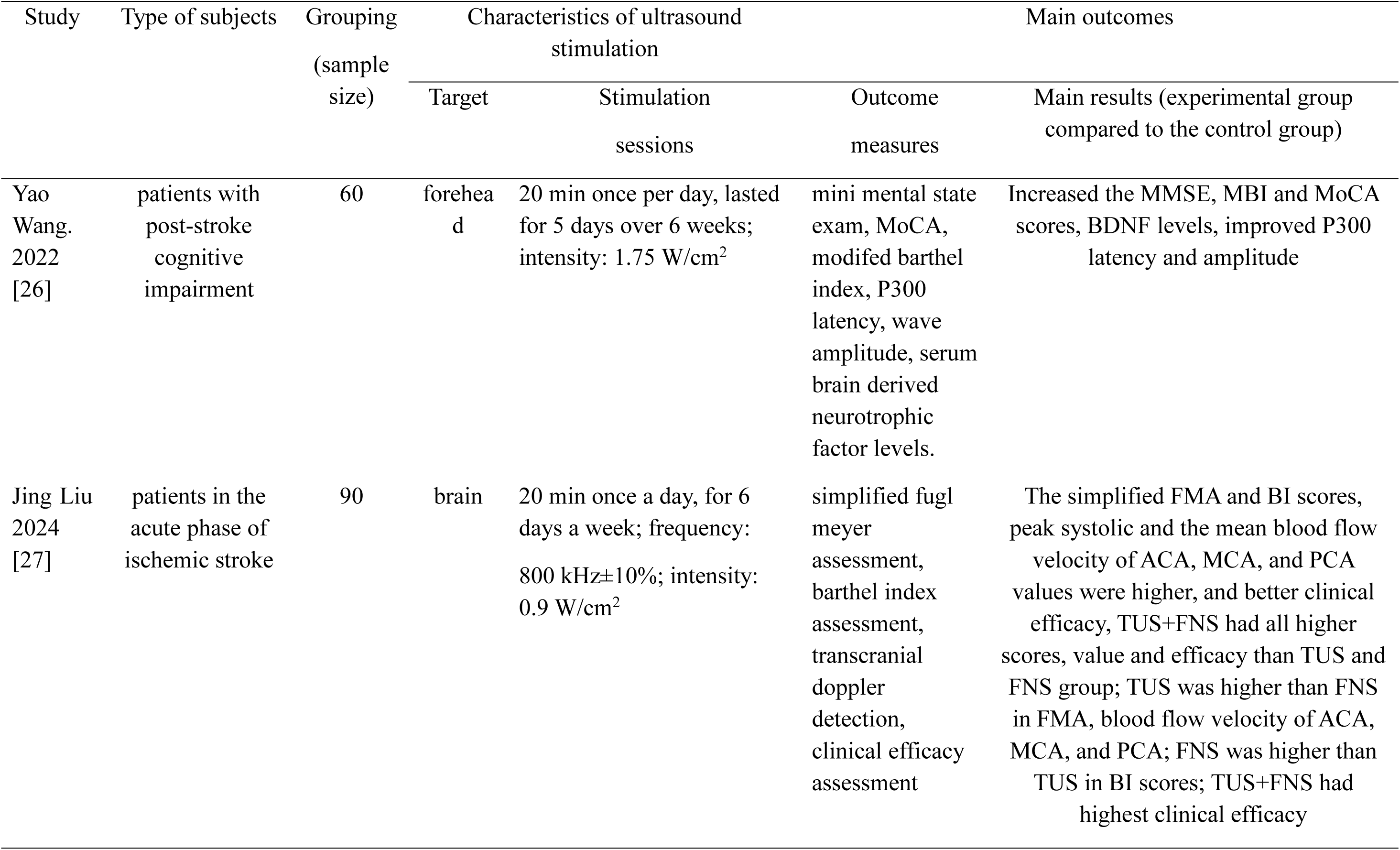
Characteristics of included human studies.

| Study | Type of subjects | Grouping<br>(sample size) | Characteristics of ultrasound stimulation |  |  | Main outcomes |
| --- | --- | --- | --- | --- | --- | --- |
|  |  |  | Target | Stimulation sessions | Outcome measures | Main results (experimental group compared to the control group) |
| Mengfei Zhang. 2022 [15] | healthy participants | 20 | brain | 10 min; Power: 1.2 W/cm<sup>2</sup><br>frequency: 0.8 MHz | tapping score, motor evoked potential | The tapping score was higher ; MEP latency was shorter; MEP amplitude increased |
| Yi Zhang. 2021 [21] | male healthy participants | 24 | left primary motor cortex | 15min; spatial peak temporal average intensity: 0.142 W/cm<sup>2</sup>; spatial peak pulse average intensity: 2.846 W/cm<sup>2</sup>; fundamental frequency: 500 kHz | single-pulsed transcranial magnetic stimulation-evoked motor evoked potential, stop-signal task | rTUS increased MEP amplitudes, improved motor inhibitory control ability |
| Nardin Samuel. 2022 [9] | healthy participants | 15 | left motor cortex | train of pulses: 80s; pulse repetition frequency: 5 Hz, pulse duration: 20 ms, ultrasonic stimulus duration: 200 ms, duty cycle: 10%, total number: 400 pulses; power of ultrasound: 20W | single-pulse TMS, paired-pulse TMS, electromyography (EMG) recording | tbTUS increased MEP amplitudes, decreases SICI, but did not alter ICF, impacted alpha and beta spectral power in distinct regions |
|  |  |  | Target | Stimulation sessions | Outcome measures | Main results (experimental group compared to the control group) |
| Yao Wang. 2022 [26] | patients with post-stroke cognitive impairment | 60 | forehead | 20 min once per day, lasted for 5 days over 6 weeks; intensity: 1.75 W/cm<sup>2</sup> | mini mental state exam, MoCA, modified barthel index, P300 latency, wave amplitude, serum brain derived neurotrophic factor levels. | Increased the MMSE, MBI and MoCA scores, BDNF levels, improved P300 latency and amplitude |
| Jing Liu 2024 [27] | patients in the acute phase of ischemic stroke | 90 | brain | 20 min once a day, for 6 days a week; frequency: 800 kHz±10%; intensity: 0.9 W/cm<sup>2</sup> | simplified fugl meyer assessment, barthel index assessment, transcranial doppler detection, clinical efficacy assessment | The simplified FMA and BI scores, peak systolic and the mean blood flow velocity of ACA, MCA, and PCA values were higher, and better clinical efficacy, TUS+FNS had all higher scores, value and efficacy than TUS and FNS group; TUS was higher than FNS in FMA, blood flow velocity of ACA, MCA, and PCA; FNS was higher than TUS in BI scores; TUS+FNS had highest clinical efficacy |

### 3.2 Methodological quality

All 18 studies were analyzed and detailed scores were listed in Table 3 and Table 4, with Table 3 shown the animal studies and Table 4 shown the human studies, respectively. Figure 2 presented the risk of bias of five human studies with no studies classified to have a high risk of bias. The overall results were that both animal and human studies had relatively good qualities.

**Table 3.** Methodological quality of the included animal studies.

| Items/reference numbers | Eligibility criteria specified | Random allocation | Comparable baseline | Assessor blinding | Clear exclusion statement | Both sexes | Less than 15% dropouts | Between group comparison | Point estimates and variability | Clear treatment protocol | Clear numbers in groups | Total score (0–10) |
| --- | --- | --- | --- | --- | --- | --- | --- | --- | --- | --- | --- | --- |
| S I.2021[16] | + | 1 | 1 | 1 | 1 | 0 | 0 | 1 | 1 | 1 | 1 | 8 |
| L L.2019[17] | + | 1 | 1 | 1 | 0 | 0 | ? | 1 | 1 | 1 | 1 | 7 |
| S C.2016[18] | + | 1 | 1 | 0 | 0 | 0 | ? | 1 | 1 | 1 | 1 | 6 |
| H M.2015[19] | + | ? | 1 | 0 | 0 | 0 | ? | 1 | 1 | 1 | 0 | 4 |
| L L.2018[20] | + | 1 | 1 | 1 | 0 | 0 | ? | 1 | 1 | 1 | 1 | 7 |
| L Q.2024[22] | + | 1 | 1 | 1 | 0 | 0 | ? | 1 | 1 | 1 | 1 | 7 |
| H B.2018[23] | + | 1 | 1 | 0 | 0 | 0 | ? | 1 | 1 | 1 | 1 | 6 |
| R C.2023[8] | + | 1 | 1 | 0 | 1 | 0 | ? | 1 | 1 | 1 | 1 | 7 |
| H B.2020[24] | + | 1 | 1 | 0 | 0 | 0 | ? | 1 | 1 | 1 | 1 | 6 |
| T G.2015[25] | + | 1 | 1 | 1 | 0 | 0 | ? | 1 | 1 | 1 | 1 | 7 |
| J W.2020[28] | + | ? | 1 | 1 | 0 | 0 | ? | 1 | 1 | 1 | 1 | 6 |
| L D.2020[29] | + | 1 | 1 | 1 | 0 | 0 | ? | 1 | 1 | 1 | 0 | 6 |
| E K.2021[30] | + | 1 | 1 | 1 | 0 | 0 | 1 | 1 | 1 | 1 | 1 | 8 |

**Table 4.**
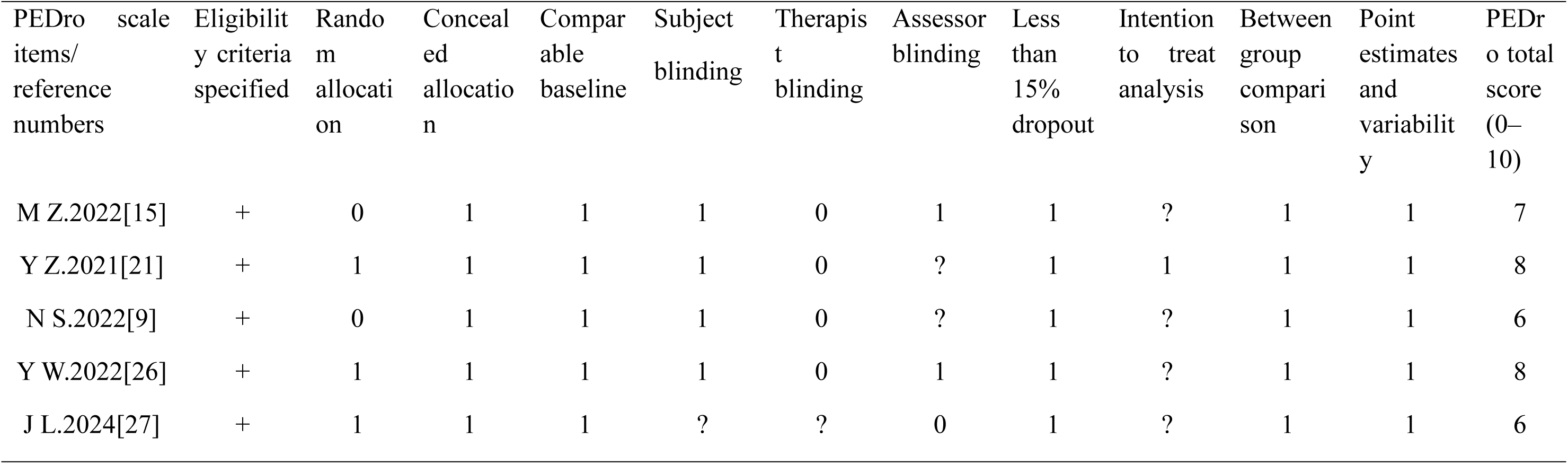
Methodological quality of the included human studies.

**Figure 2.**
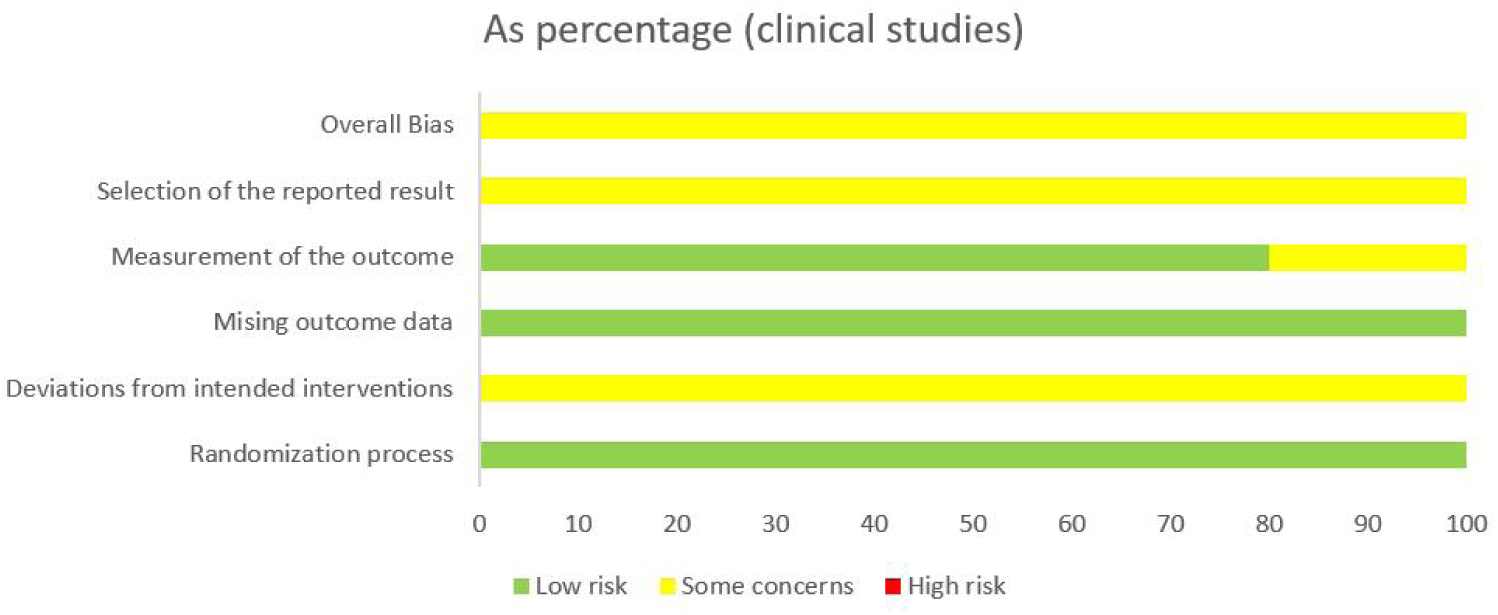
Risk of bias of included human studies

There was one animal studies that contained supply form to show the dropouts [16], in addition that another study presented at least in one test, it has less than 15% dropouts [30]. Two animal studies didn’t emphasize whether they randomly allocated mice in groups [19][28], and two human studies didn’t divide participants into groups [15][9]. In addition, that all animal tests were carried out on male mice. In Yi Zhang’s paper [21], it mentioned in what condition data would be excluded, while it also mentioned no participant was excluded, so we decided to give it 1 score in “Intention to treat analysis”.

### 3.3 Effects of transcranial ultrasound stimulation on stroke recovery

The effects of TUS on stroke recovery were categorized under three domains: clinical and pre-clinical, neuroimaging and neurophysiological, and biochemical outcomes.

The main outcomes were listed in Table 1 and Table 2 and more detailed outcomes would be discussed in the following sub-chapters.

There were some examinations for the tables. MCAO was a commonly applied method to produce stroke models in mice, which was the abbreviation of middle cerebral artery occlusion. It was a method for inducing focal cerebral ischemia [11], in addition that distal MCAO [25] and transient MCAO [30] could be induced in different ways. After the experiment, different evaluation methods would be applied to prove the effectiveness of the therapy. Observe and scoring behaviors among different groups was a widely used technique to evaluate the outcomes of one therapy on stroke recovery, which might include distinct methods between different subjects [15]. In addition, different types of dyes will be applied to tissue and introduce visible color to injury tissues, vessels, and cells, PCR can be used for RNA detection and Weston blot is for proteins [16]. Besides these, some physical methods will also be applied. For example, laser speckle contrast imaging can monitor the change of cerebral blood flow [8].

#### 3.3.1 Clinical and pre-clinical outcomes

There were a variety of clinical and pre-clinical outcomes on stroke rehabilitation that were illustrated by TUS, while in this study, motor functions and cognitive functions would be mainly discussed.

Among all animal studies, different behavior tests were carried out to evaluate the effects of TUS. There were totally 10 studies that did these kinds of tests, and all illustrated that ultrasound stimulation contributed to motor function recovery. The stimulation was applied to the stroke region or the injury hemisphere, and after therapy, moving speed, body balance, walking distance and other changes could be observed. While there was one study [16] figured out that there were no significant differences between control group and stimulation group in corner test and Barnes circular maze test.

There were three human studies that tested the motor functions of participants, and one tested the cognitive functions, with mini mental state exam (MMSE), modified Barthel index (MBI) and MoCA were applied for cognitive effects [26]. The results showed that if the TUS was combined with other therapies, like fastigial nucleus stimulation (FNS) treatment [27] or conventional cognitive rehabilitation training [26], it would gain better improvement compared to applying TUS or other therapy alone. In addition, if the TUS was applied alone, it would still have better outcomes than FNS group [27]. While it was reported that after TUS, the accuracy of body movement was not improved [21], it only shortened the reaction time.

#### 3.3.2 Neuroimaging and neurophysiological outcomes

In this section, outcomes mainly tested on physical methods would be listed. For example, the change of blood flow, brain edema, delta activity, cerebral hemodynamic, apparent diffusion coefficient (ADC), motor evoked potential (MEP), short interval intracortical inhibition (SICI), intracortical facilitation (ICF), alpha and beta spectral power, P300 latency and amplitude, the peak systolic blood flow velocity (Vs) and the mean blood flow velocity (Vm).

The overall outcome was that TUS could significantly improve the above indexes. It was worth noting that in one animal study, TUS suppressed the pathological delta activity, which was high right after stroke occurrence, and resulted in restoring interhemispheric balance [24]. In addition, one animal study only reported the change in cerebral hemodynamic [30]. While there were two animal studies reported that if the TUS therapy was applied 9 hours after stroke occurrence, it would not affect the ADC [17][20], which meant TUS wouldn’t have curative effect on acute cerebral infarction after 9 hours’ delay.

In human studies, there was one study reported that ICF wasn’t greatly alerted by TUS [9], in addition, another study found that TUS didn’t significantly change the MEP latency [21].

What’s more, there was one study combining TUS with FNS, and the results were that TUS+FNS group would have higher values of Vs and Vm than both the TUS and FNS groups, respectively [27], and TUS group gain higher values than FNS group. The corresponding velocities were detected in anterior cerebral artery (ACA), middle cerebral artery (MCA), and posterior cerebral artery (PCA).

#### 3.3.3 Biochemical outcomes

Among all outcomes, there were a large number of outcomes that were examined through biochemical tests, and they would be listed below. They mainly included the change of the injury area, the number of neural related cells and vessels, the expression of neural related proteins and RNA, the brain blood barrier and the serum brain-derived neurotrophic factor (BDNF) levels.

Among all 13 animal studies, there were 9 papers that reported the change of the injury area, and most of the findings were that TUS would decrease the injury area and protect the brain. While there was one study that reported TUS didn’t differ in the infarct volume among control groups and experimental groups [29], and another only applied the infarct area as evidence of successfully making the stroke model [20].

There were 4 animal studies presenting TUS could increase the number of positive vessels or microvessels, and one study showed that TUS promoted the remodeling of leptomeningeal vasculature and the growth of vessels in length along with density [8]. TUS was also effective in cell growth. In total 7 papers mentioned TUS could reduce neuronal degeneration and neuronal loss [17], increased mesenchymal stem cells (MSC) and neural-like cells migration to the wound area [18], decreased the number of neutrophils that leading to less inflammation in the brain [25] and promoted the growth and number of other neural related cells.

The gene expression is always related to the change of RNA and proteins. In total 4 paper tested the change of RNA and 7 paper did the protein test. The overall findings were that TUS adjusted gene expression, which resulted in an increase or decrease of certain types of RNA or protein and contributed to stroke recovery. In addition, more detailed mechanisms of TUS effect on blood flow recovery and newly formed vessels were found, the related genes were also listed and discussed [22].

There was one animal study reported that TUS could attenuate blood brain barrier disruption [29], and one human study showed that BDNF levels in the TUS group significantly increased [26].

### 3.4 Effectiveness—active stimulation versus sham

The data included in this part was divided into three parts: tissue, excitability and motor function. In this part, not all indexes mentioned above were included, for some of them didn’t have enough data for analysis. In addition, that the cognitive functions would not be discussed in this section, for only one paper present it. We couldn’t find enough evidence for further analysis. The “tissue” part included all indexes of tissue level, for example, the content of RNA, certain kind of cell and proteins and the wound area. The “excitability” part contained indexes of organ or system level, like MEP and blood flow. In total, the post-interventional data indicated the effect of TUS on the parameters. Tables 5, 6 and 7 presented the effect sizes of all three subgroups, and Figures 3, 4, and 5 showed the overall effect size. Fisher’s method was applied for all three subgroups, and the results were listed in Table 8, with degree of freedom of each group.

**Table 5.**
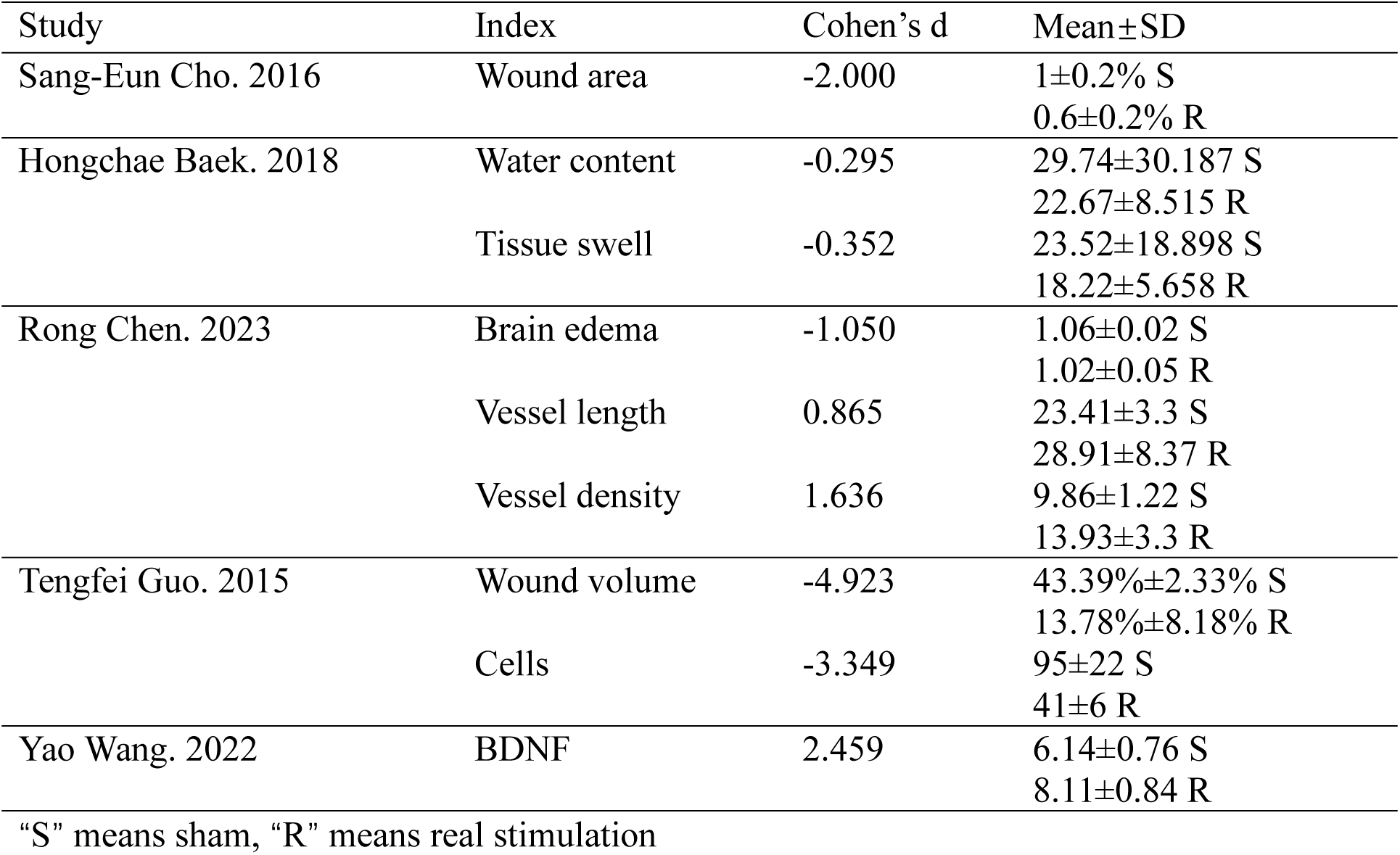
Tissue.

| Study | Index | Cohen's d | Mean±SD |
| --- | --- | --- | --- |
| Sang-Eun Cho. 2016 | Wound area | -2.000 | 1±0.2% S<br>0.6±0.2% R |
| Hongchae Baek. 2018 | Water content | -0.295 | 29.74±30.187 S<br>22.67±8.515 R |
|  | Tissue swell | -0.352 | 23.52±18.898 S<br>18.22±5.658 R |
| Rong Chen. 2023 | Brain edema | -1.050 | 1.06±0.02 S<br>1.02±0.05 R |
|  | Vessel length | 0.865 | 23.41±3.3 S<br>28.91±8.37 R |
|  | Vessel density | 1.636 | 9.86±1.22 S<br>13.93±3.3 R |
| Tengfei Guo. 2015 | Wound volume | -4.923 | 43.39%±2.33% S<br>13.78%±8.18% R |
|  | Cells | -3.349 | 95±22 S<br>41±6 R |
| Yao Wang. 2022 | BDNF | 2.459 | 6.14±0.76 S<br>8.11±0.84 R |

“S” means sham, “R” means real stimulation

**Table 6.**
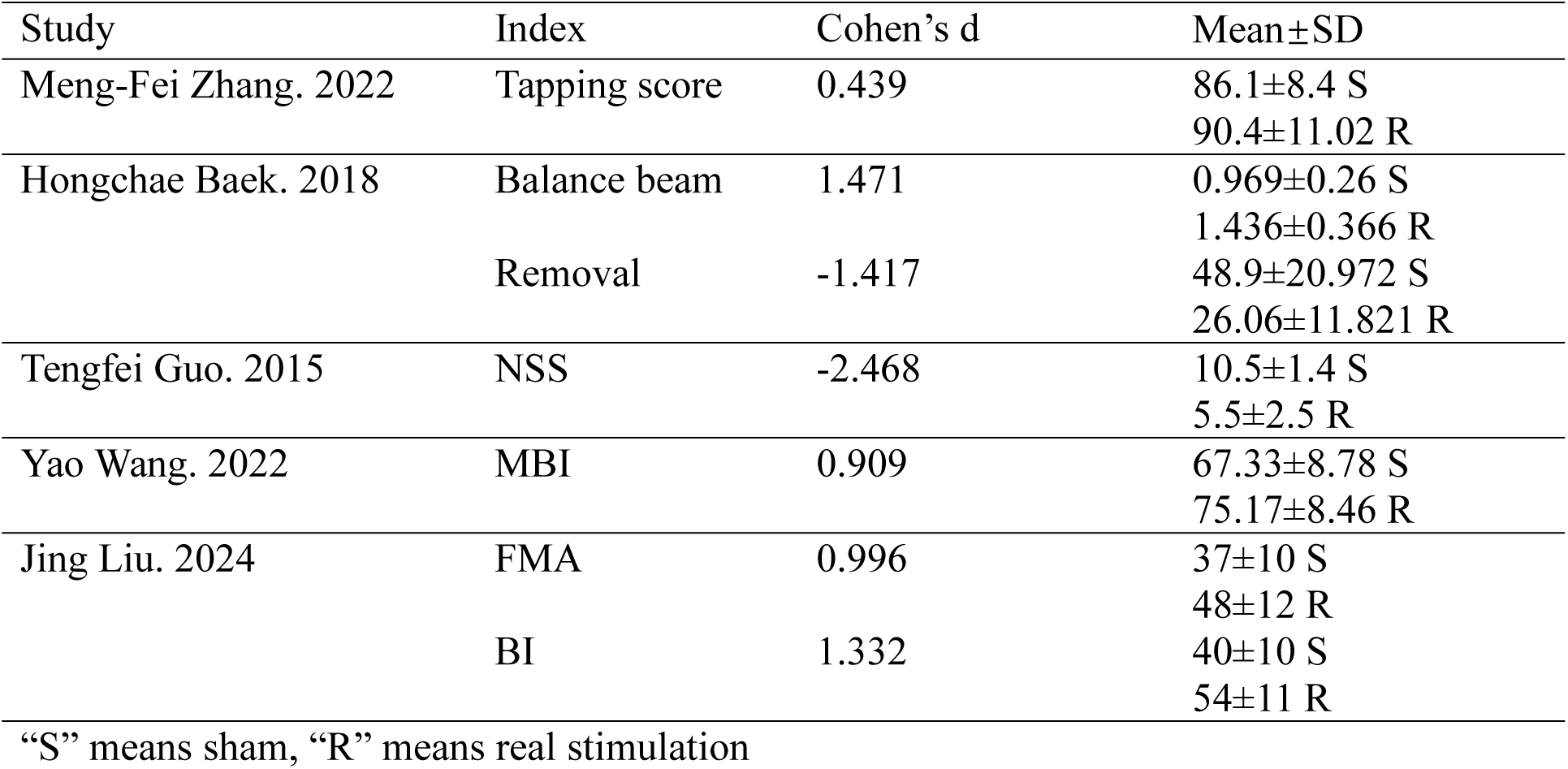
Motor functions.

| Study | Index | Cohen's d | Mean±SD |
| --- | --- | --- | --- |
| Meng-Fei Zhang. 2022 | Tapping score | 0.439 | 86.1±8.4 S<br>90.4±11.02 R |
| Hongchae Baek. 2018 | Balance beam | 1.471 | 0.969±0.26 S<br>1.436±0.366 R |
|  | Removal | -1.417 | 48.9±20.972 S<br>26.06±11.821 R |
| Tengfei Guo. 2015 | NSS | -2.468 | 10.5±1.4 S<br>5.5±2.5 R |
| Yao Wang. 2022 | MBI | 0.909 | 67.33±8.78 S<br>75.17±8.46 R |
| Jing Liu. 2024 | FMA | 0.996 | 37±10 S<br>48±12 R |
|  | BI | 1.332 | 40±10 S<br>54±11 R |

“S” means sham, “R” means real stimulation

**Table 7.**
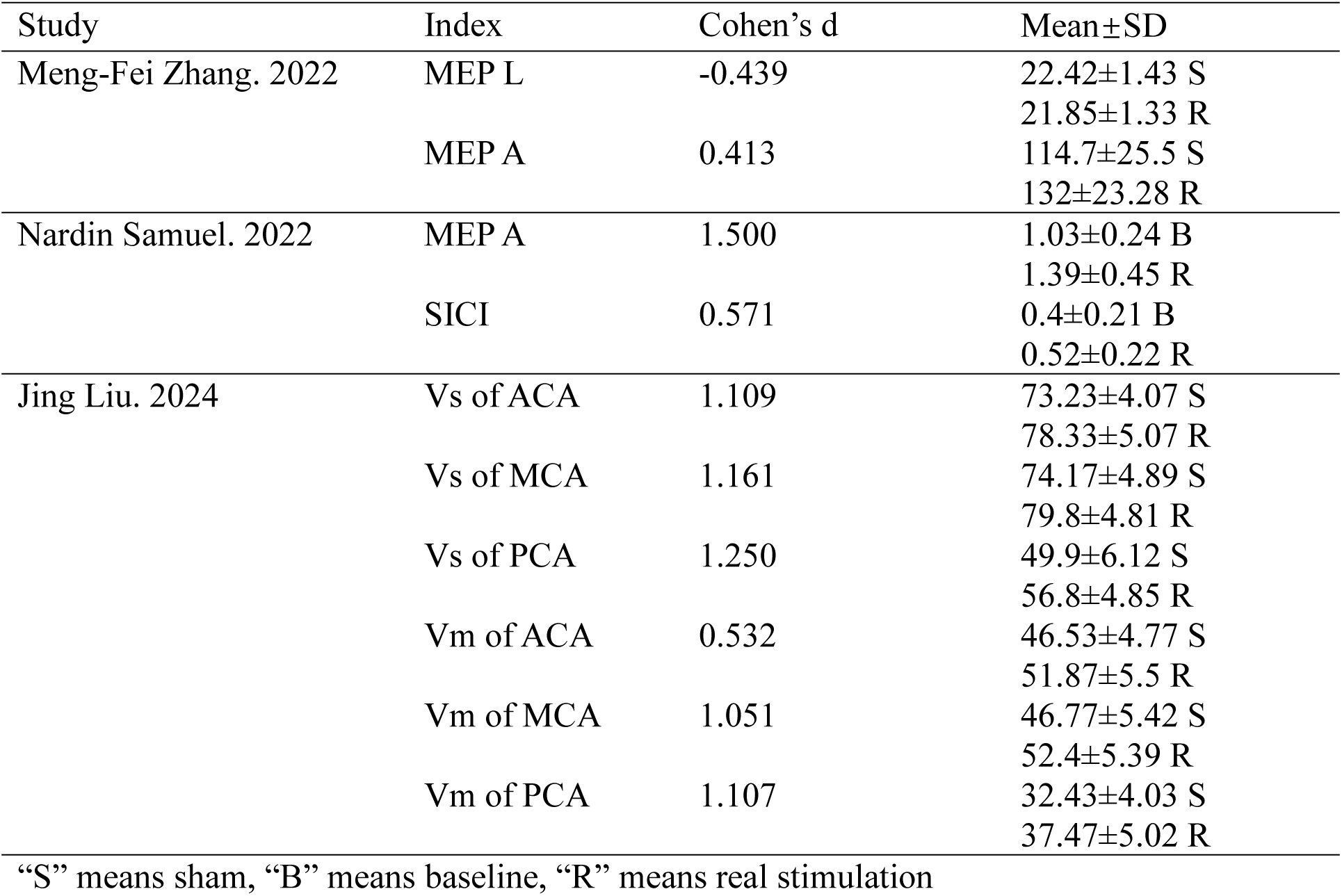
Excitability.

| Study | Index | Cohen's d | Mean±SD |
| --- | --- | --- | --- |
| Meng-Fei Zhang. 2022 | MEP L | -0.439 | 22.42±1.43 S<br>21.85±1.33 R |
|  | MEP A | 0.413 | 114.7±25.5 S<br>132±23.28 R |
| Nardin Samuel. 2022 | MEP A | 1.500 | 1.03±0.24 B<br>1.39±0.45 R |
|  | SICI | 0.571 | 0.4±0.21 B<br>0.52±0.22 R |
| Jing Liu. 2024 | Vs of ACA | 1.109 | 73.23±4.07 S<br>78.33±5.07 R |
|  | Vs of MCA | 1.161 | 74.17±4.89 S<br>79.8±4.81 R |
|  | Vs of PCA | 1.250 | 49.9±6.12 S<br>56.8±4.85 R |
|  | Vm of ACA | 0.532 | 46.53±4.77 S<br>51.87±5.5 R |
|  | Vm of MCA | 1.051 | 46.77±5.42 S<br>52.4±5.39 R |
|  | Vm of PCA | 1.107 | 32.43±4.03 S<br>37.47±5.02 R |

“S” means sham, “B” means baseline, “R” means real stimulation

**Figure 3.**
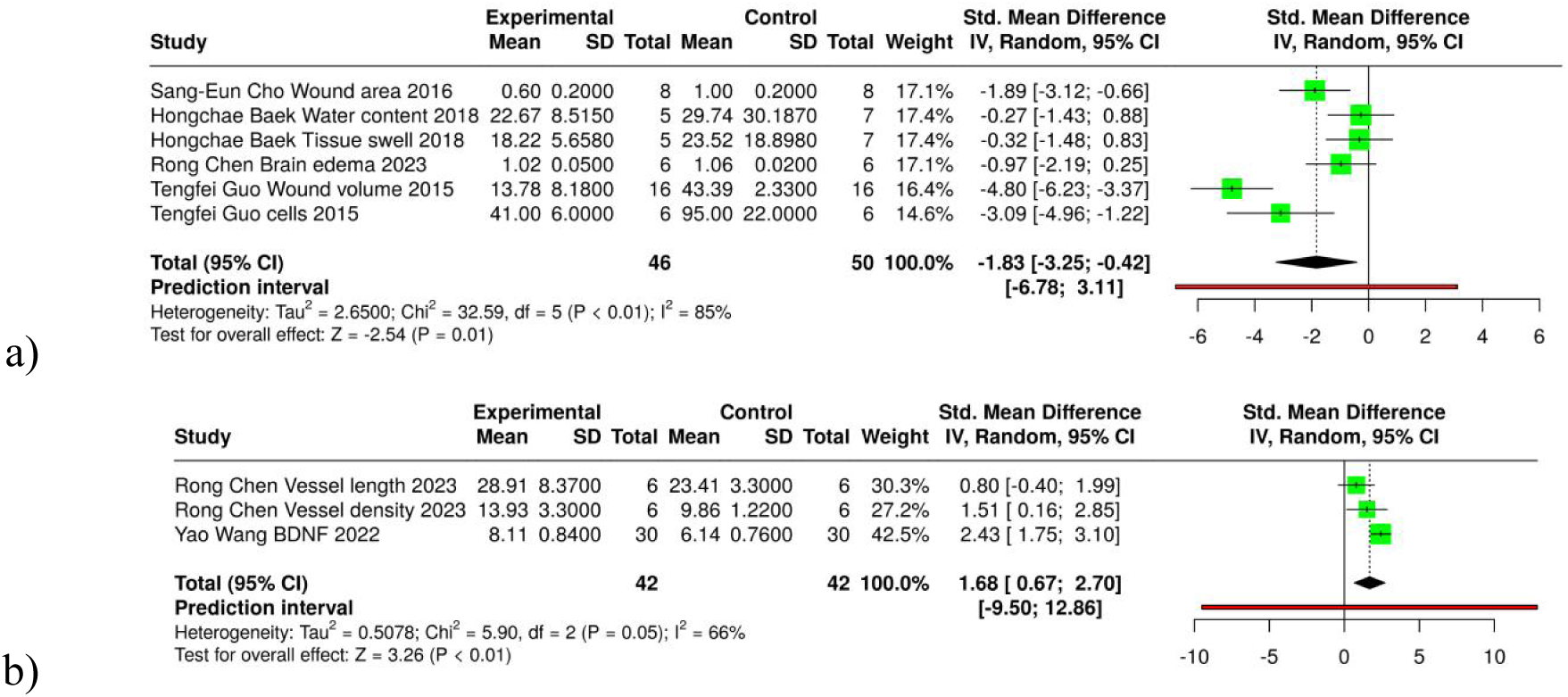
Forest plot depicting the effect of TUS on tissue domain. “Water content” and “tissue swell” were both related to the wound are recovery. “Cells” referred to numbers of neutrophils. The figure a) referred to the negative changes and b) indicated the positive change

**Figure 4.**
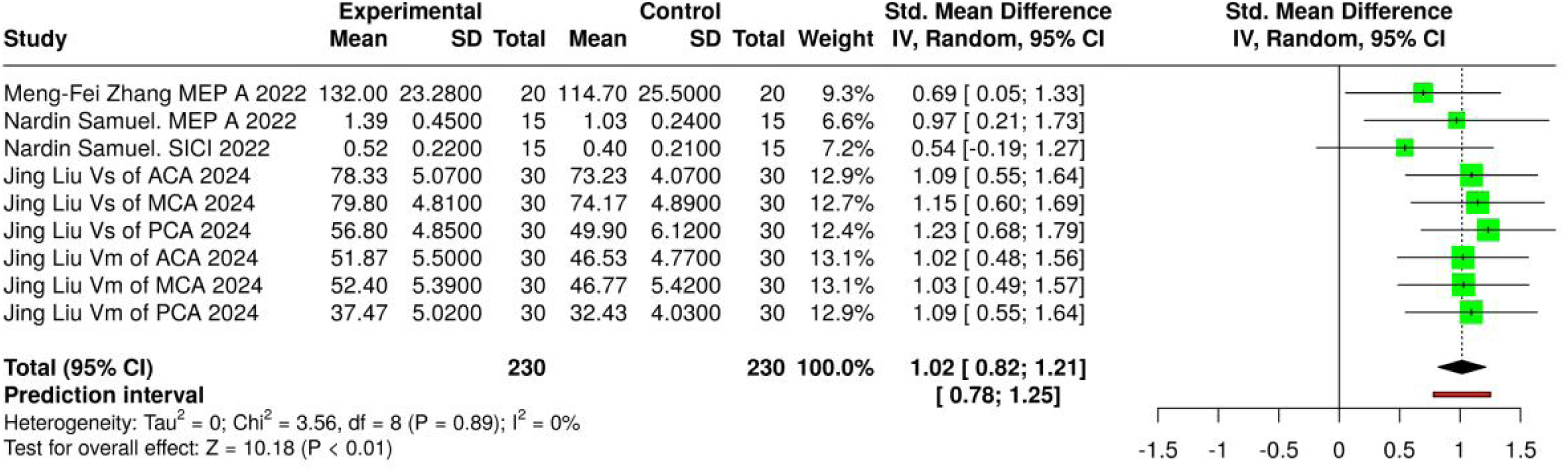
Forest plot depicting the effect of TUS on excitability domain. “MEP A” referred to motor evoked potential amplitude, respectively. “SICI” meant short interval intracortical inhibition, which could indicate motor cortical excitability. “Vs” and “Vm” referred to peak systolic blood flow velocity and mean blood flow velocity respectively, with “ACA” meant anterior cerebral artery, “MCA” referred to middle cerebral artery and “PCA” was the abbreviation of posterior cerebral artery

**Figure 5.**
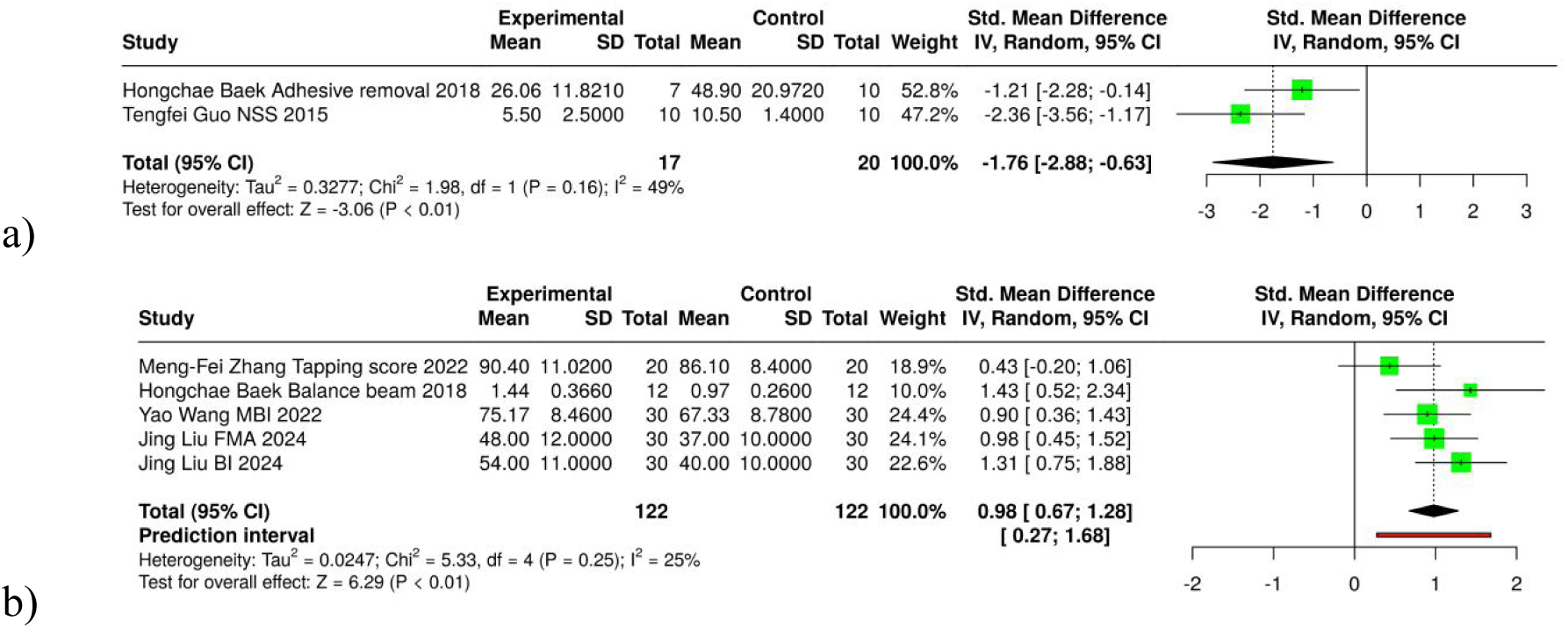
Forest plot depicting the effect of TUS on motor function domain. “NSS” was the abbreviation of neurological severity score indicating the motor functions. “MBI” was modified Barthel index, measuring an individual’s level of independence in activities of daily living. “FMA” and “BI” referred to fugl-meyer assessment and Barthel index assessment respectively, and both were related to motor functions. The figure a) referred to the negative changes and b) indicated the positive change

**Table 8.** Fisher’s method.

| Subgroup | Fisher's X2 | Degree of freedom |
| --- | --- | --- |
| Excitability | 192.4 | 40 |
| Tissue | 458.7 | 122 |
| Motor | 191.6 | 48 |

There were some indexes reduced after the treatment and led to a negative number in the table, these also meant the stimulation had positive effects on stroke recovery, which included the reduce of wound area and brain edema, the decrease of water content and tissue swelling and less immune responses reflected by less neutrophils. In addition, if the adhesive removal time and NSS decreased, the movements of mice with stroke models were more close to healthy objects.

## 4. Discussion

In this review, we comprehensively summarized the effects of TUS on clinical and pre-clinical, neurophysiological, neuroimaging, and biochemical outcomes in human and animal research. TUS enhanced motor function, blood flow, blood flow velocity and other indexes in healthy subjects and stroke models, improved cognitive function, and protected brain tissue. It also adjusted gene expression, reduced inflammatory response and promoted the rebuilding of injury tissue. No histological, biochemical, or behavioral side effects were identified in the animal or human studies included in this review.

### 4.1 Safety

Among all the included studies, most papers applied certain TUS protocol for experiments. It has evidence that the previous studies had already established some reliable therapeutic frameworks for TUS, which included safe dosage of acoustic waves and the treatment parameters [31]. In animal studies, previous studies found that, if using focused TUS, the minimum intensities for motor responses were: 4.9–5.6 W/cm^2^ in spatial peak pulse average intensity, and 2.5–2.8 W/cm^2^ in spatial peak temporal average intensity, or 1–5 ms of tone-burst duration, 50% of duty cycle, and 300 ms of duration, at 350 kHz fundamental frequency [32]. While the reality is that different stimulation methods may have slightly different protocol, for example, unfocused TUS might need higher intensity to reach the same outcome. When it comes to human studies, the choices of protocols are always referring to some previous animal tests, and the lowest effective stimulation intensity should be higher because of the skull thickness [33], and the lowest intensity is related to different people. Based on current knowledge, it’s suggested that before clinical trials, perform some simulation studies and in vitro studies to determine the appropriate dosage. Besides the dosage, therapy duration is another important parameter, for TUS may cause heat-related damage and mechanical damage [34]. Although more detailed studies are not found, because the safe duration is related to many parameters mentioned above, FDA recommends that MI should be less than 1.9, which is an estimation of the risk [35]. The target selection is of great importance, for the brain has highly region-specific nature [36]. Most animal tests included in this review choose to stimulate the injured region or the same hemisphere. While for human studies, the target choice is more general. The chosen regions were usually related to some certain functions, and some studies simply applied the TUS to the entire brain.

Considering that between animal models and human objects, the energy transfer rate of tissue and the skull thickness are different, which might enable intenser or more powerful stimulation to be applied on clinical trails. While on clinical trails, current imaging based regulatory pathways may limit this application. The limitations for TUS given by the International Consortium for Transcranial Ultrasonic Stimulation Safety and Standards (ITRUSST) are: 1) the MI or the MI_tc_ does not exceed 1.9 and 2) a temperature rise less than 2℃, a thermal dose less than 0.25 CEM43, or specific values of the TI for a given exposure time [44]. The above indexes of a stimulation protocol can be estimated and calculated through certain formula. For example, the temperature rise can be calculated using absorption coefficient, ultrasound intensity, ultrasound stimulation duration, the density of brain tissue, and the specific heat of brain tissue [25]. In Yi Zhang’s paper of their clinical experiment, the maximum temperature increase after intermittent stimulation was 6.4×10^−3^℃ in the skull and 6.1×10^−4^℃ in the brain, and after 2.65s continuous stimulation, the maximum temperature increase was 0.134℃ in the skull and 9.1×10^−3^℃ in the brain [21]. In this case, just assuming ten times of intensity is applied with other parameters unchanged, the maximum temperature rise is 1.34℃ in the skull, which should be safe. In Evgenii Kim’s preclinical test, the temperature increase was not higher than 0.14℃ and 0.0018℃ for skull and brain, respectively [30]. It can be see that the applied intensities for both clinical and preclinical trails are far below the safety limitation, which may not fully accounts for the advancements in FUS technology and its potential benefits. More specific regulatory pathways might be crucial to establish.

### 4.2 Possible mechanism

This review indicates a possible explanation for the mechanism of applying TUS to stroke recovery. Primarily, the ion channels, signal pass ways and cells related to stroke recovery are activated through cavitation, temperature or mechanical deformation [45]. These cells produce more RNA and proteins to help the injured tissue, which can lead to vessel growth and decrease of immune response. When vessels are formed, more nutrition can be carried to the wound area, and more stem cells can migrate and differentiate there. This might help the injured tissue to keep or regain some previous functions. At the same time, TUS excites different parts of cortex, restore interhemispheric communication, and accelerates the processes of function compensation. The healthy but not previously activated domains can replace the functions of damaged parts. These may both contribute to functional recovery on individuals.

### 4.3 Effectiveness and limitations

Regarding the quality assessment, most of the dropout rates in animal studies are unknown, in addition that only male mice are used, which may lead to unbalanced outcomes among studies. In addition, that most included studies didn’t mention the exclusion strategies in statistical analysis, this can be a risk for having variable effectiveness on individuals. Moreover, in human studies, most of the therapists and assessors are not blind to the grouping, which may also result in some uncontrollable outcomes.

There are in total three reports showing that TUS can be combined with other therapies, and all gained better outcomes. This might be a potential application of TUS. While up to now, only a few research has been done on this, especially for combining TUS with other transcranial stimulation methods. Thus, whether TUS can be applied together with other stimulation methods, and whether this kind of combination can have better outcomes, these still remain unknown. More research could be done to answer these questions.

When it comes to the limitation of this review, firstly, in consideration of the strict searching strategies and eligibility criteria, some potential studies of great importance might not be included in this review. Secondly, we did not search for studies that exclusively investigated some more detailed stimulation protocols and parameters of TUS. The included studies might be insufficient to assess the full effective section of TUS [14].

## 5. Conclusion

TUS is a promising non-invasive stimulation therapy that modulates brain activities in animals and humans. The effects on stroke recovery include improving motor and cognitive functions, altering gene expression and regulating other biochemical processes like inflammatory responses and vessel growth. If TUS is combined with other therapy, it will gain more significant outcomes. While the mechanisms of TUS remain unknow, and future research is recommended to investigate the optimal treatment parameters and strategies on humans.

## Data Availability

All data produced in the present study are available upon reasonable request to the authors

## Notes

### Competing Interest Statement

The authors have declared no competing interest.

### Funding Statement

This study did not receive any funding

### Author Declarations

Databases like PubMed, Web of Science and Scopus

